# Implementation Assessment of an Integrated Early Childhood Development and Violence Prevention Program: Evidence from Conflict-Affected Antioquia, Colombia (2013-2022)

**DOI:** 10.64898/2025.12.30.25343241

**Authors:** Alexandra Restrepo-Henao, Nilton Montoya Gomez, Javier Orlando Contreras, José William Cornejo Ochoa, Ana María Castaño, Maria Patricia Arbelaez Montoya

**Affiliations:** Professor, Violence and risk behaviors research. Epidemiology Research Group. National School of Public Health. Universidad of Antioquia. Colombia. Address: 295 N. Martin Ave. Drachman Hall; Statistician. Database specialist, Statistics and public health. National School of Public Health. Universidad of Antioquia. Colombia; Pediatrics Department, Research in Child and Adolescent Disorders (Pediaciencia). Pediatrics Department. Universidad of Antioquia., Colombia; Advisor National Council of Accreditation Colombia CNA, Neurologist, Pediatric Neurology, MSc Epidemiology. Professor. Research in Child and Adolescent Disorders (Pediaciencia). Pediatrics Department. Universidad of Antioquia. Colombia.; Psychologist; Emeritus professor, University of Antioquia, Colombia MPH, PhD in epidemiology

## Abstract

**Introduction:** Programs that integrate early childhood and violence prevention should be assessed for implementation, particularly in low- and middle-income countries and conflict settings. We assessed participants’ satisfaction and the implementation barriers and facilitators of a program to promote child development and prevent early aggressive behaviors from the prenatal period through 36 months in Antioquia, Colombia.

**Materials and methods:** A mixed-method design was used. A quantitative survey was conducted with child caregivers (n = 5,919) to assess caregiver satisfaction. In addition, a qualitative study assessed barriers to and facilitators of program implementation. We analyzed longitudinal fieldwork diaries collected during implementation (n=84) and semi-structured interviews with healthcare professionals (n=5) and caregivers (n=6).

**Results:** Caregivers reported 95% satisfaction across all activities. Barriers and facilitators were identified: 1) Outer settings (Context): lack of implementation of public policies and violence in communities. Some transversal categories include health system financial crisis, lack of cooperation between institutions, beliefs and attitudes toward health promotion programs, and the predominance of the biomedical paradigm over the health promotion approach. 2) Inner stings (Health service): lack of health promotion programs and financial difficulties within health services. 3) Individual characteristics: caregiver characteristics, including being a first-time mother, having prior positive experiences with health services, and sharing experiences with other caregivers through program activities. Healthcare worker characteristics: lack of training in health promotion activities and precarious working conditions. 4) Program: number of activities, online training, adaptation, and theoretical support.

**Conclusions:** Barriers to implementing a program to promote child development and prevent early aggressive behavior included living in low-income communities, financial instability in the health system, limited cooperation among health institutions, poor working conditions for healthcare workers, and beliefs about prevention approaches.

## Introduction

Researchers and practitioners advocated for the integration of Early prevention violence strategies and Early Child Development Programs (ECDP)(1). This integration could create a stimulating environment for the children that supports optimal development and reduces early violence (2). The implementation of these programs has been essential for children and families in low-income settings and in conflict or post-conflict areas, as it can mitigate the impact of these stressors on child development (3). While ECDP has proven effective (4, 5), few studies have integrated ECDP with violence prevention (6). Furthermore, the effectiveness can be undermined by diverse barriers when implemented in the ‘natural’ conditions of health services. The gap between program design and program delivery could represent high costs for health systems and families who do not receive the high-quality services required to promote children’s mental health (2). Implementation research can contribute to understanding implementation gaps by systematically studying the characteristics of interventions, processes, and contextual factors that determine the intervention’s efficacy (7). However, research on the determinants of the implementation of child development programs in both high- and low- and middle-income countries (LMIC) is scarce.

A systematic review assesses the implementation pathways and strategies of ECDP in LMIC. They identified 75 articles representing 33 programmers across 23 LMICs. They describe how collaboration across multiple sectors supports program success, the program’s adaptation to communities, and the staff, training, and coaching during the implementation. They highlight the importance of context in implementation and how the lack of infrastructure, differences in community needs, and cultures can influence it,(8).

However, few studies address implementation gaps when integrating the early violence prevention programs into ECDP. Black et al. conducted a hybrid design in Rwanda (778 caregivers and 55 children) to assess the integration of ECDP plus violence prevention. They trained non-specialists to implement the program through home-visit strategies. The program showed effectiveness in stimulating children at home, caregiving involvement, reducing harsh discipline, and increasing dietary diversity. The program also rated the outcomes as moderate across different implementations (9).

The current evidence reported the importance of the *outer and inner setting*s, highlighting how constant changes in the leadership made by a policy maker, lack of leadership and cooperation between health institutions, working conditions of healthcare workers, deficits in the program design and planning, caregiver geographical access, and a user’s previous experiences with health services as determinants of implementation barriers.

The outer setting plays a crucial role in the implementation of health promotion policies. However, policymakers’ frequent turnover, the lack of institutional leadership and interprogram cooperation, and the working conditions of healthcare workers have been described as barriers to ECDP implementation (10, 11). Often, when a policymaker changes positions or officers, they may alter entire programs, policies, staff, procedures, and funding mechanisms. In Australia, the Triple P program was scaled up, benefiting approximately 10,000 children (12). During implementation, researchers found that turnover of politicians and health professionals impedes program continuity and affects access to funding over time. Therefore, researchers and program implementers are forced to develop constant communication strategies to engage with new stakeholders (12).

Within the inner setting, Sanders et al. highlight the leadership of healthcare workers and how cooperation between and within institutions, as well as the participation of diverse stakeholders in program design, facilitates implementation. To successfully implement the programs, it is necessary to have strong leadership in health services that follows the program’s orientation and supports cooperation between institutions (12).

In addition, the Family Education and Support Program (FESP) from Spain (10) and the Maternal and Child Health and Nutrition Program (MCHNP) from Argentina found that the lack of clear leadership among child development healthcare workers often led to implementing different procedures and reducing fidelity (11). Similarly, the lack of cooperation between professionals and health institutions reduces program fidelity and increases costs, as professionals create parallel activities and pursue different goals and procedures (10, 11).

Regarding the individual characteristics, some studies show that the importance of the healthcare worker’s individual factors, dissatisfaction with work, and stress could interfere with program implementation and quality. The evaluation of MCHNP found that providers experience high stress levels due to their work in low-income communities (11). Family needs outweighed professional and health services responsibilities, thereby creating hopelessness among healthcare workers. Also, researchers found that healthcare workers’ jobs are unstable but demanding. This can be an obstacle to implementation because, under these working conditions, the program was perceived as an additional burden, thereby diminishing healthcare workers’ interest in program quality and continuity (11).

Individual characteristics, such as limited transportation resources and prior experiences with health services, and caregiver characteristics may determine parents’ engagement in the program. The MCHNP reports that geographic proximity of an institution to a caregiver’s home facilitates participation in the program’s activities. Additionally, research describes the history of contact with healthcare services, and satisfaction with those services may determine engagement in new programs and adherence to their activities. Parents who detect prior fragmentation in healthcare services and the lack of service articulation are less likely to engage in early interventions. Caregivers in the MCHNP found that professionals were not articulate and repeated the same information, thereby reducing parents’ satisfaction and engagement with the program (11). Some processes are important facilitators of program implementation. Program characteristics related to design and planning are critical to success. The program’s length, the number of activities, and the balance in the number of participants per activity are essential facilitators of program efficacy. FESP reported that high fidelity, an average of 10 activities, and a short program length increased the program’s fidelity (10). However, there should be a minimum program dose (i.e., the number and duration of activities) to achieve the desired effect (10).

Similarly, both small and large caregiver groups can face difficulties in interacting and sharing experiences. Programs are successful when they provide training and technical support to healthcare workers, because implementing a new intervention entails behavioral changes; therefore, healthcare workers must learn and internalize the latest practices to maintain program fidelity (10).

This article describes the integration of a program designed to promote child development and prevent early aggressive behavior into the Grow and Developmental program implemented in Colombia’s health services. We evaluate satisfaction levels, barriers, and facilitators of programs that promote child development and prevent early aggressive behavior from the prenatal period through 36 months in Antioquia, Colombia. To assess the implementation determinants, we used the Consolidated Framework for Implementation Research (CFIR)(13), which has been proposed to understand the program implementation process and outcomes. In this model, the determinants of the implementation are the outer settings, inner settings, individual factors, process innovation, and implementation process. We conducted surveys to evaluate caregivers’ satisfaction, and collected qualitative longitudinal data from multiple actors (i.e., policymakers, healthcare workers, and caregivers). These results not only provide evidence to improve PBFP implementation but also yield essential insights into the facilitators and barriers to successful implementation of similar programs in LMICs. These findings may also provide important information about how to implement ECDP in low-income communities and conflict conditions, increasing the evidence about the implementation of ECDP (2, 14).

### Materials and methods Study design

Using the multilevel implementation research approach, we conducted a mixed-methods evaluation of program implementation. We assessed the parents’ and healthcare workers’ satisfaction with the program through a quantitative descriptive study, and we evaluated implementation barriers and facilitators using a qualitative research based on content analysis (15)

### Intervention

The Promise of a Better Future Program (PBFP) was designed to promote child development and prevent early aggressive behaviors by drawing on evidence on how to support child development in low-income settings. The program was adapted and implemented between December 1^st,^ 2015, and December 15^th,^ 2021. This program was adapted and incorporated into growth and development programs due to its high prestige within communities, and it was offered free to insured and uninsured individuals, as well as to low-income communities. In addition, hospitals and municipalities in the public sector allocated personnel and resources to this program.

We conducted quantitative and qualitative components to evaluate the implementation presented in the current analysis. This analysis is part of a broad evaluation that includes a cluster-randomized control trial. We invited 26 municipalities and public health institutions in Antioquia state, including the Western, Eastern, and Metropolitan regions.

The program was based on four main theories and evidence: 1) the theory of child development model proposed by Bronfenbrenner (16), 2) the trans-theoretical model of change (17), 3) review of the evidence about the effectiveness of different programs to prevent child aggression and promote child development, and 4) review of the evidence about the relationship between positive parenting practices and child development and aggressive behaviors.

In graphic 1, we can observe the components of the PBFP, and we explain below: *1) Program adaptation and improvement:* the PBFP was designed and improved according to caregiver and service needs. After, we conducted a series of meetings and interviews with stakeholders and families to improve and adapt the program. *2) Dissemination:* the PBFP was disseminated through a diploma course (the diploma course was called Promes@) and communicational activities. The training aimed to strengthen healthcare workers’ knowledge and skills to promote child development and prevent early aggressive behaviors. Trained healthcare workers implemented the program in the communities. The training has two components: a) 72 hours of theoretical training through an internet-based course, and b) 88 hours of practical activities implemented in the program. The program was disseminated through community activities and communication channels, including radio and TV programs, videos, social media posts, and other community initiatives. *3) Program implementation:* The third component of the program was implementing PBFP in the health services. Trained healthcare workers implemented the program in communities during practicum placements. The program comprises 27 individual or group activities with children’s caregivers. These activities began during the last trimester of pregnancy and continued monthly through the first two years of life. The program was designed to conduct activities either in groups within health institutions or with individuals in caregivers’ homes, depending on community security conditions. Before the pandemic, the activities were group activities at health institutions or individual activities at children’s homes. However, during the COVID-19 pandemic, we adapted the intervention by creating a virtual version. We developed a new intervention manual for conducting virtual activities. *4) Evaluation:* the program evaluation includes the implementation’s barriers and strengths that are presented in this article and a Cluster-RCT.

### Quantitative study about caregivers’ satisfaction and pertinence

We conducted a descriptive study to describe caregivers’ satisfaction with the program. Caregivers’ dissatisfaction with the program can be a significant barrier to implementation. The study participants were PBFP members who had participated in program activities. The inclusion and exclusion criteria are summarized in Table 1.

**Table 1.**
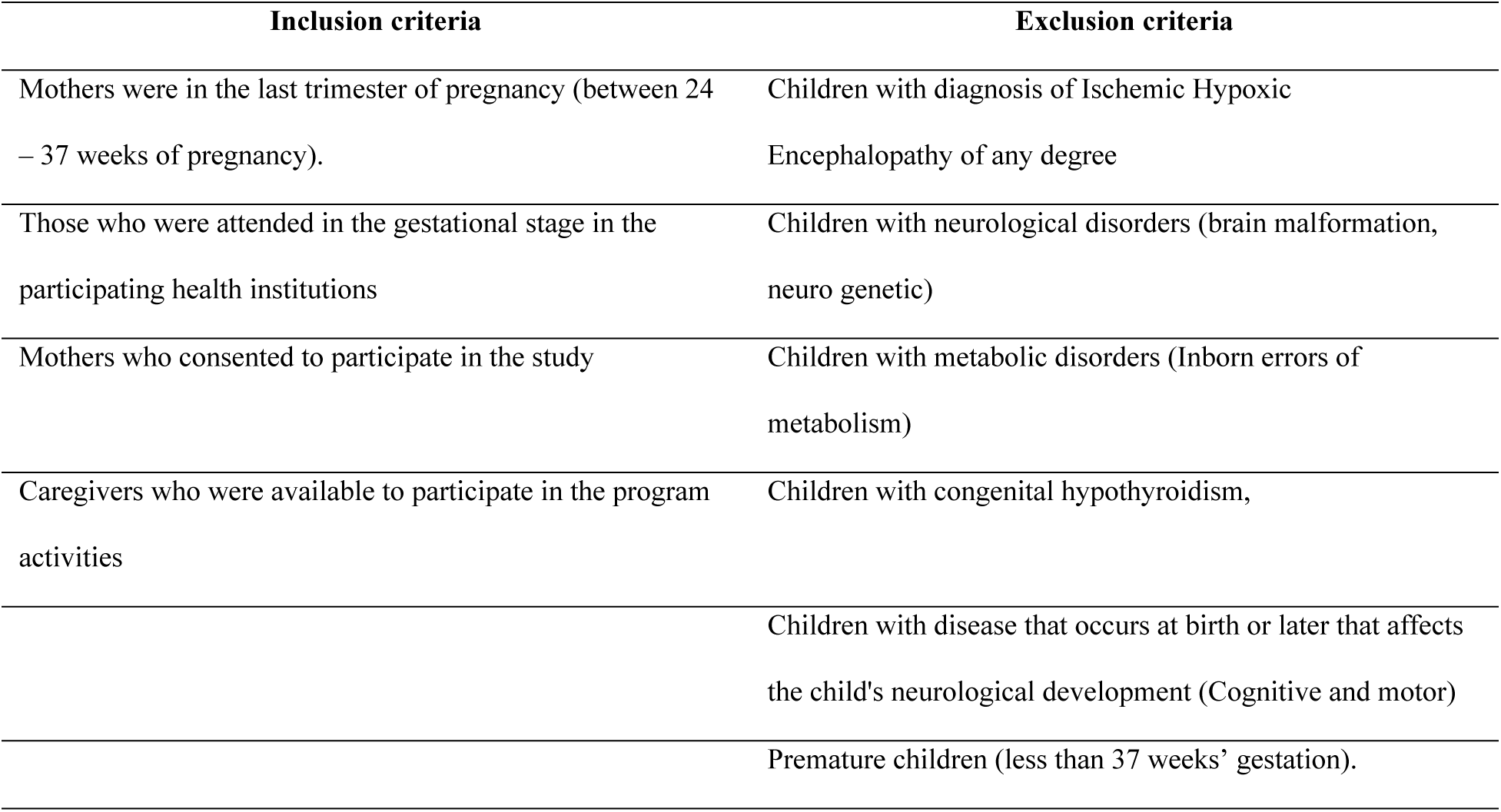
Inclusion and exclusion criteria for participation in Promise of Better Future Program.

We collected satisfaction surveys immediately after each activity with the caregiver. This survey was anonymous and self-administered (n=579 caregivers; 5,919 satisfaction surveys). We employed a team-specific satisfaction questionnaire adapted from a similar instrument used in a program for schoolchildren (18). This questionnaire comprised 12 items assessing satisfaction with the activity, its facilitators, duration, location, and relevance. We calculated a score that ranged from 1 to 100. In the analysis, we present descriptive information about caregivers and children. We then presented the mean and standard deviation of satisfaction for each satisfaction activity, stratified by demographic variables (i.e., gender, age, socioeconomic level, and first-time mothers versus mothers with more than one child). To compare satisfaction means, we used a two-sample t-test when the satisfaction score had a normal distribution, and a Mann-Whitney U Test when the satisfaction score did not follow a normal distribution. We calculated Kruskal-Wallis’ test to assess the mean difference across mothers’ education levels.

### Qualitative component

We conducted a quantitative study to understand the barriers and facilitators to implementation from the perspectives of different actors. We collected qualitative information using three strategies: 1) fieldwork diaries: each fieldwork professional had a fieldwork diary on paper or in a computer, in which they wrote their impression and experiences working with the healthcare workers, observing the activities performed by the healthcare workers, and meeting with the hospital and municipality teams. Those diaries were collected on paper and transcribed for a research assistant. 2) Opportunities, weaknesses, and threats report (SWOT): We also conducted periodical SWOT reports (n=82) using a participatory approach with the healthcare workers and the leadership in each hospital. We held monthly meetings with healthcare workers and each municipality’s point person to discuss barriers and facilitators and to improve the program. 3) semi-structured interviews: At the end of the third year, we collected five semi-structured interviews with professionals and six caregivers. We conducted interviews with professionals and caregivers who participated in the overall process. We invited the point professionals from each municipality, and five accepted to participate. Interviews were coded and transcribed from the audio-recordings. In addition, we invited all caregivers to participate in the qualitative study, and six mothers agreed to participate.

For the qualitative data analysis, we developed an initial codebook based on the constructs identified in prior research. Two investigators conducted the coding; the procedure was standardized. Emergent categories were discussed, defined, and incorporated into the codebook using the five-factor model (7). The first coding was conducted with the fieldwork diaries and reports. Researchers developed focus group questions based on the initial coding to address gaps in those categories with insufficient information. The initial codes were merged into sub-categories that shared similar content discussed between researchers. The coding was performed in Excel 2016.

## Results

### Caregiver and family characteristics

Table 2 presents selected characteristics of mothers and families who participated in the program activities. The program included 579 caregivers who attended 5,919 activities over the past four years. The mean gestational age at study entry was 30.56 months, and the mean maternal age was 25 years. Among families, 80% were from low socioeconomic strata; 29% of mothers had completed elementary school, and 45% had completed high school. In addition, 82% of mothers were housewives, while 1% were working, and 68% had the government subsidize health insurance (Table 2).

**Table 2.**
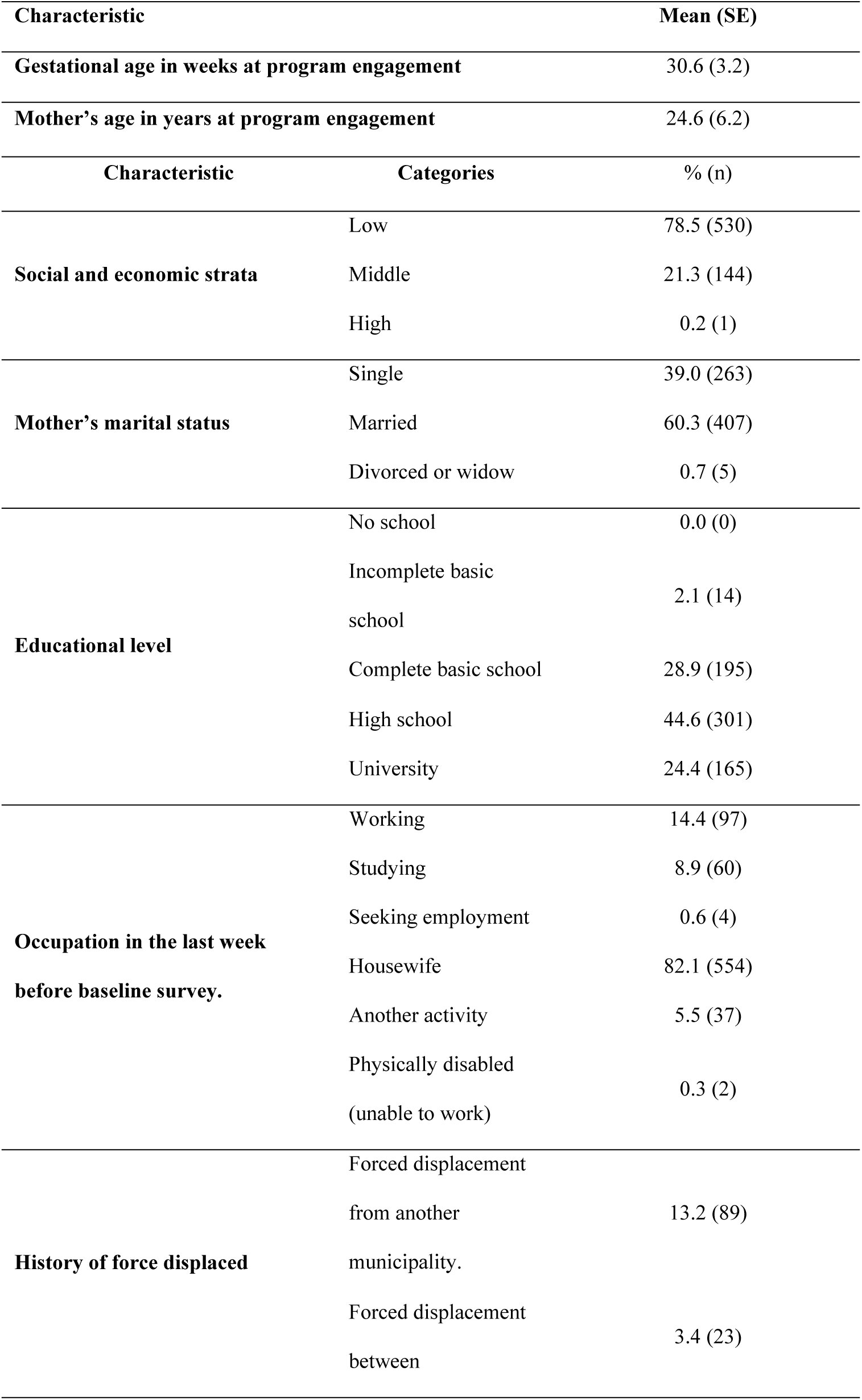
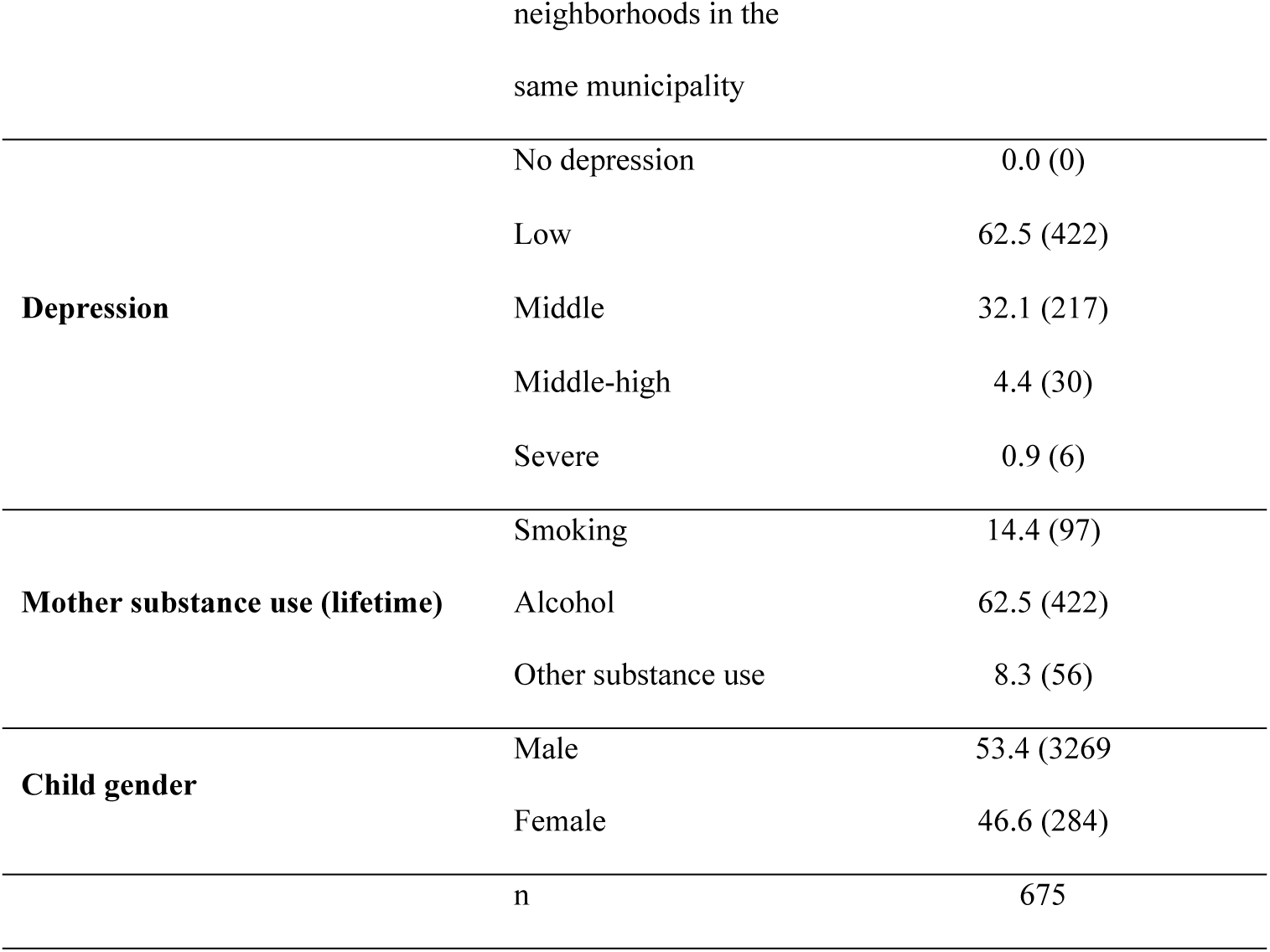
The Promise of Better Future Program participant characteristics.

Regarding maternal mental health, we found that 4% of mothers had middle-high level of depression, and 68% consumed alcohol in their lifetime. Of children’s families, 13% were forcefully displaced by illegal armed groups (Table 2).

### Caregiver’s satisfaction: quantitative component

During the study period, we conducted 5,376 caregiver satisfaction evaluations. The caregiver satisfaction with the program was high for all activities (greater than 95%) (See Table 3). In general, mothers’ caregiver satisfaction was similar across demographic variables, except for satisfaction with motivation, life project, and caregiver self-care activities, which were lower for those caregivers with a complete basic education. Among caregiver’s suggestions, we found that 31% of caregivers reported they would want better spaces to perform the activities, 15% would prefer to improve the adherence to the PBFP, 15% provided innovative activities such as hydrotherapy and physical activity, 9% suggested the program should have more activities, and 5% performed more dynamic activities (See Table 4).

**Table 3.**
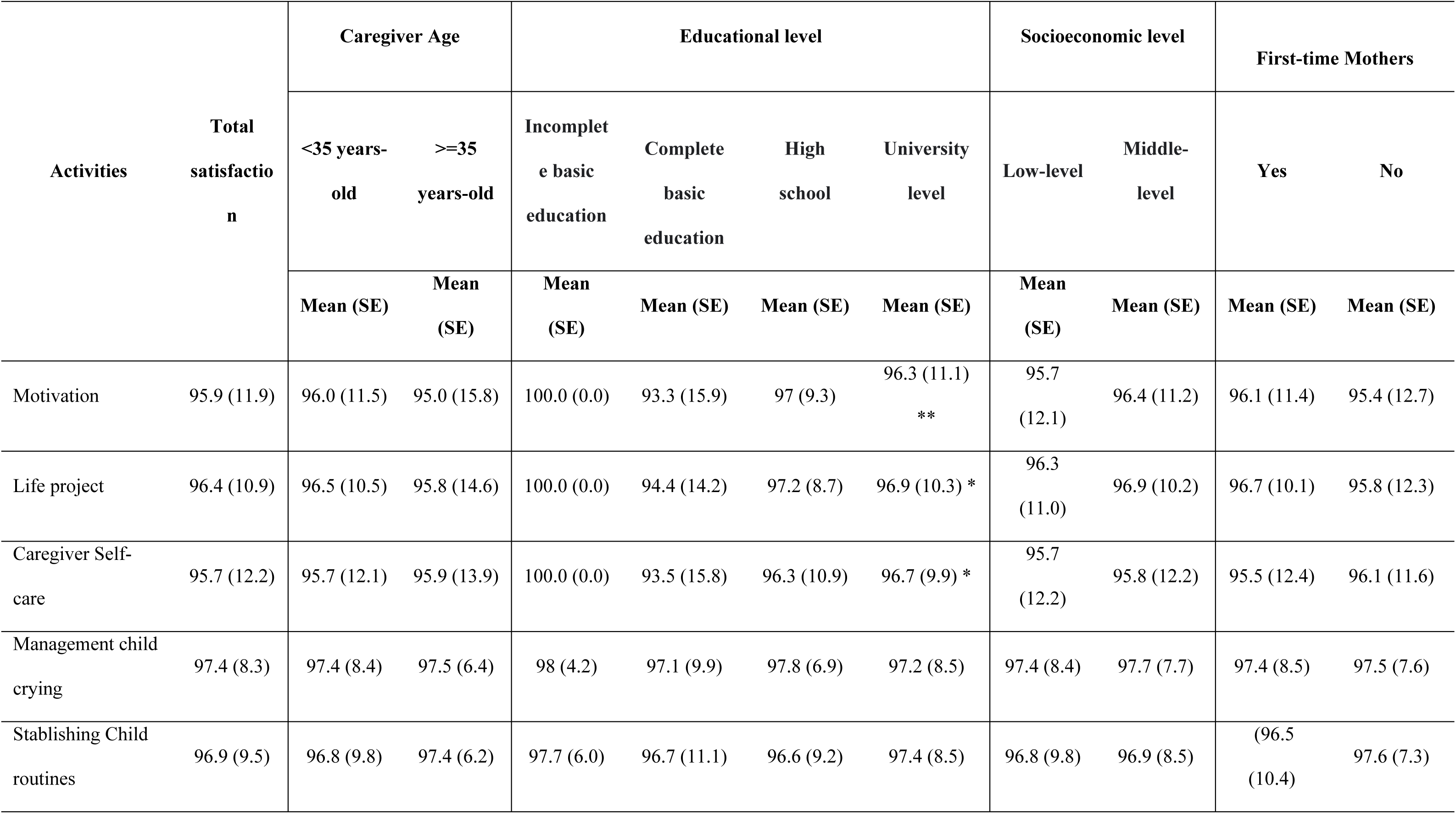
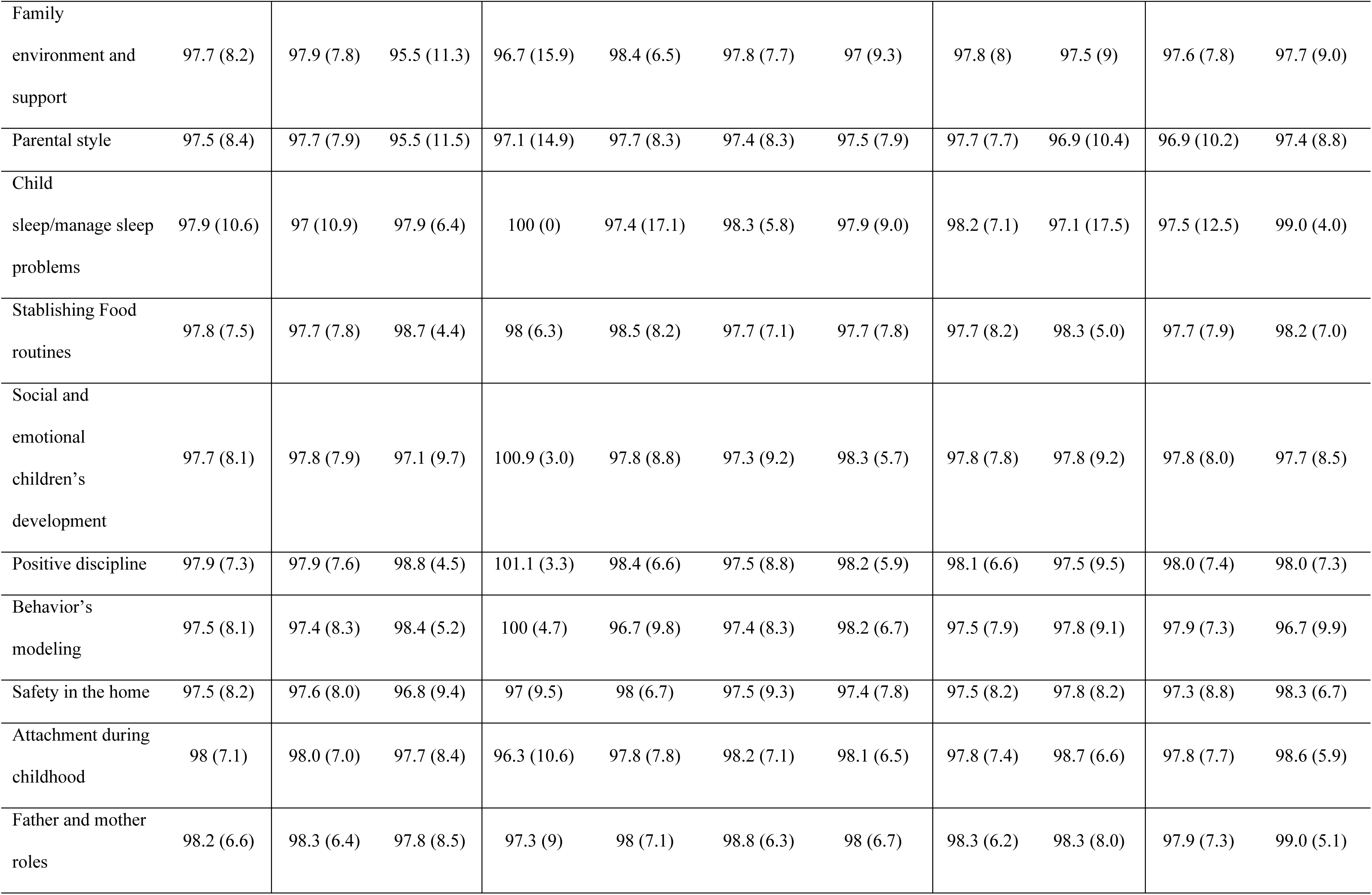
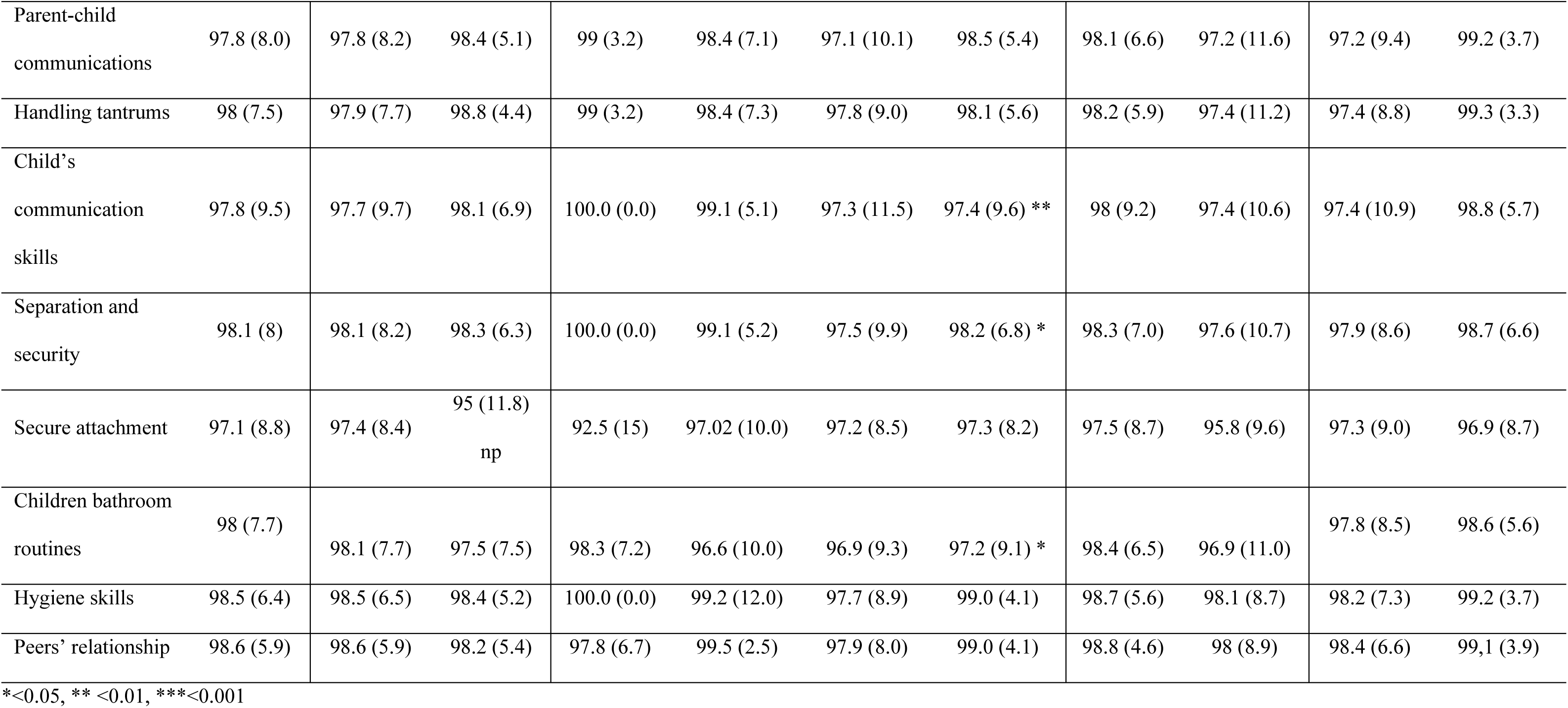
Caregiver’s satisfaction score per program activity, mothers’ age, mother educational level, socioeconomic level and first-time mothers (5,919 satisfaction surveys to 579 participants).

| Activities | Total satisfaction | Caregiver Age |  | Educational level |  |  |  | Socioeconomic level |  | First-time Mothers |  |
| --- | --- | --- | --- | --- | --- | --- | --- | --- | --- | --- | --- |
|  |  | <35 years-old | >=35 years-old | Incomplete basic education | Complete basic education | High school | University level | Low-level | Middle-level | Yes | No |
|  |  | Mean (SE) | Mean (SE) | Mean (SE) | Mean (SE) | Mean (SE) | Mean (SE) | Mean (SE) | Mean (SE) | Mean (SE) | Mean (SE) |
| Motivation | 95.9 (11.9) | 96.0 (11.5) | 95.0 (15.8) | 100.0 (0.0) | 93.3 (15.9) | 97 (9.3) | 96.3 (11.1)<br>** | 95.7 (12.1) | 96.4 (11.2) | 96.1 (11.4) | 95.4 (12.7) |
| Life project | 96.4 (10.9) | 96.5 (10.5) | 95.8 (14.6) | 100.0 (0.0) | 94.4 (14.2) | 97.2 (8.7) | 96.9 (10.3) * | 96.3 (11.0) | 96.9 (10.2) | 96.7 (10.1) | 95.8 (12.3) |
| Caregiver Self-care | 95.7 (12.2) | 95.7 (12.1) | 95.9 (13.9) | 100.0 (0.0) | 93.5 (15.8) | 96.3 (10.9) | 96.7 (9.9) * | 95.7 (12.2) | 95.8 (12.2) | 95.5 (12.4) | 96.1 (11.6) |
| Management child crying | 97.4 (8.3) | 97.4 (8.4) | 97.5 (6.4) | 98 (4.2) | 97.1 (9.9) | 97.8 (6.9) | 97.2 (8.5) | 97.4 (8.4) | 97.7 (7.7) | 97.4 (8.5) | 97.5 (7.6) |
| Stablishing Child routines | 96.9 (9.5) | 96.8 (9.8) | 97.4 (6.2) | 97.7 (6.0) | 96.7 (11.1) | 96.6 (9.2) | 97.4 (8.5) | 96.8 (9.8) | 96.9 (8.5) | (96.5 (10.4) | 97.6 (7.3) |
| Family environment and support | 97.7 (8.2) | 97.9 (7.8) | 95.5 (11.3) | 96.7 (15.9) | 98.4 (6.5) | 97.8 (7.7) | 97 (9.3) | 97.8 (8) | 97.5 (9) | 97.6 (7.8) | 97.7 (9.0) |
| Parental style | 97.5 (8.4) | 97.7 (7.9) | 95.5 (11.5) | 97.1 (14.9) | 97.7 (8.3) | 97.4 (8.3) | 97.5 (7.9) | 97.7 (7.7) | 96.9 (10.4) | 96.9 (10.2) | 97.4 (8.8) |
| Child sleep/manage sleep problems | 97.9 (10.6) | 97 (10.9) | 97.9 (6.4) | 100 (0) | 97.4 (17.1) | 98.3 (5.8) | 97.9 (9.0) | 98.2 (7.1) | 97.1 (17.5) | 97.5 (12.5) | 99.0 (4.0) |
| Stablishing Food routines | 97.8 (7.5) | 97.7 (7.8) | 98.7 (4.4) | 98 (6.3) | 98.5 (8.2) | 97.7 (7.1) | 97.7 (7.8) | 97.7 (8.2) | 98.3 (5.0) | 97.7 (7.9) | 98.2 (7.0) |
| Social and emotional children's development | 97.7 (8.1) | 97.8 (7.9) | 97.1 (9.7) | 100.9 (3.0) | 97.8 (8.8) | 97.3 (9.2) | 98.3 (5.7) | 97.8 (7.8) | 97.8 (9.2) | 97.8 (8.0) | 97.7 (8.5) |
| Positive discipline | 97.9 (7.3) | 97.9 (7.6) | 98.8 (4.5) | 101.1 (3.3) | 98.4 (6.6) | 97.5 (8.8) | 98.2 (5.9) | 98.1 (6.6) | 97.5 (9.5) | 98.0 (7.4) | 98.0 (7.3) |
| Behavior's modeling | 97.5 (8.1) | 97.4 (8.3) | 98.4 (5.2) | 100 (4.7) | 96.7 (9.8) | 97.4 (8.3) | 98.2 (6.7) | 97.5 (7.9) | 97.8 (9.1) | 97.9 (7.3) | 96.7 (9.9) |
| Safety in the home | 97.5 (8.2) | 97.6 (8.0) | 96.8 (9.4) | 97 (9.5) | 98 (6.7) | 97.5 (9.3) | 97.4 (7.8) | 97.5 (8.2) | 97.8 (8.2) | 97.3 (8.8) | 98.3 (6.7) |
| Attachment during childhood | 98 (7.1) | 98.0 (7.0) | 97.7 (8.4) | 96.3 (10.6) | 97.8 (7.8) | 98.2 (7.1) | 98.1 (6.5) | 97.8 (7.4) | 98.7 (6.6) | 97.8 (7.7) | 98.6 (5.9) |
| Father and mother roles | 98.2 (6.6) | 98.3 (6.4) | 97.8 (8.5) | 97.3 (9) | 98 (7.1) | 98.8 (6.3) | 98 (6.7) | 98.3 (6.2) | 98.3 (8.0) | 97.9 (7.3) | 99.0 (5.1) |
| Parent-child communications | 97.8 (8.0) | 97.8 (8.2) | 98.4 (5.1) | 99 (3.2) | 98.4 (7.1) | 97.1 (10.1) | 98.5 (5.4) | 98.1 (6.6) | 97.2 (11.6) | 97.2 (9.4) | 99.2 (3.7) |
| Handling tantrums | 98 (7.5) | 97.9 (7.7) | 98.8 (4.4) | 99 (3.2) | 98.4 (7.3) | 97.8 (9.0) | 98.1 (5.6) | 98.2 (5.9) | 97.4 (11.2) | 97.4 (8.8) | 99.3 (3.3) |
| Child's communication skills | 97.8 (9.5) | 97.7 (9.7) | 98.1 (6.9) | 100.0 (0.0) | 99.1 (5.1) | 97.3 (11.5) | 97.4 (9.6) ** | 98 (9.2) | 97.4 (10.6) | 97.4 (10.9) | 98.8 (5.7) |
| Separation and security | 98.1 (8) | 98.1 (8.2) | 98.3 (6.3) | 100.0 (0.0) | 99.1 (5.2) | 97.5 (9.9) | 98.2 (6.8) * | 98.3 (7.0) | 97.6 (10.7) | 97.9 (8.6) | 98.7 (6.6) |
| Secure attachment | 97.1 (8.8) | 97.4 (8.4) | 95 (11.8)<br>np | 92.5 (15) | 97.02 (10.0) | 97.2 (8.5) | 97.3 (8.2) | 97.5 (8.7) | 95.8 (9.6) | 97.3 (9.0) | 96.9 (8.7) |
| Children bathroom routines | 98 (7.7) | 98.1 (7.7) | 97.5 (7.5) | 98.3 (7.2) | 96.6 (10.0) | 96.9 (9.3) | 97.2 (9.1) * | 98.4 (6.5) | 96.9 (11.0) | 97.8 (8.5) | 98.6 (5.6) |
| Hygiene skills | 98.5 (6.4) | 98.5 (6.5) | 98.4 (5.2) | 100.0 (0.0) | 99.2 (12.0) | 97.7 (8.9) | 99.0 (4.1) | 98.7 (5.6) | 98.1 (8.7) | 98.2 (7.3) | 99.2 (3.7) |
| Peers' relationship | 98.6 (5.9) | 98.6 (5.9) | 98.2 (5.4) | 97.8 (6.7) | 99.5 (2.5) | 97.9 (8.0) | 99.0 (4.1) | 98.8 (4.6) | 98 (8.9) | 98.4 (6.6) | 99.1 (3.9) |

\*<0.05, \*\* <0.01, \*\*\*<0.001

**Table 4.** Caregiver’s recommendations to improve the Promise of Better Future Program.

| **Recommendations** | **%** |
| --- | --- |
| Perform playful activities with children | **5.8** |
| Improve the space where activities are performed | **30.8** |
| Provide more activities performed on time | **3.5** |
| Perform more dynamic activities | **5.1** |
| Provide longer activities | **6.1** |
| Provide economic support | **1.6** |
| Improve the family adherence | **14.1** |
| Provide more activities overtime | **9,0** |
| Schedule activities in multiple hours | **5.4** |
| Provide support from medical specialist | **3.5** |
| Provide innovative activities such as hydrotherapy<br>and physical activity | **15.1** |

### Barriers and facilitators: qualitative component

In the qualitative component, six caregivers were interviewed from different municipalities. Additionally, we interviewed five health professionals, including nurses, auxiliary nurses, and doctors. The healthcare professional’s interviewers were women (see Figure 2). Barriers and facilitators during implementations were described at different levels. The results are summarized below, and the supplement table 1 synthesizes each category and provides quotes for each category.

**Figure.**
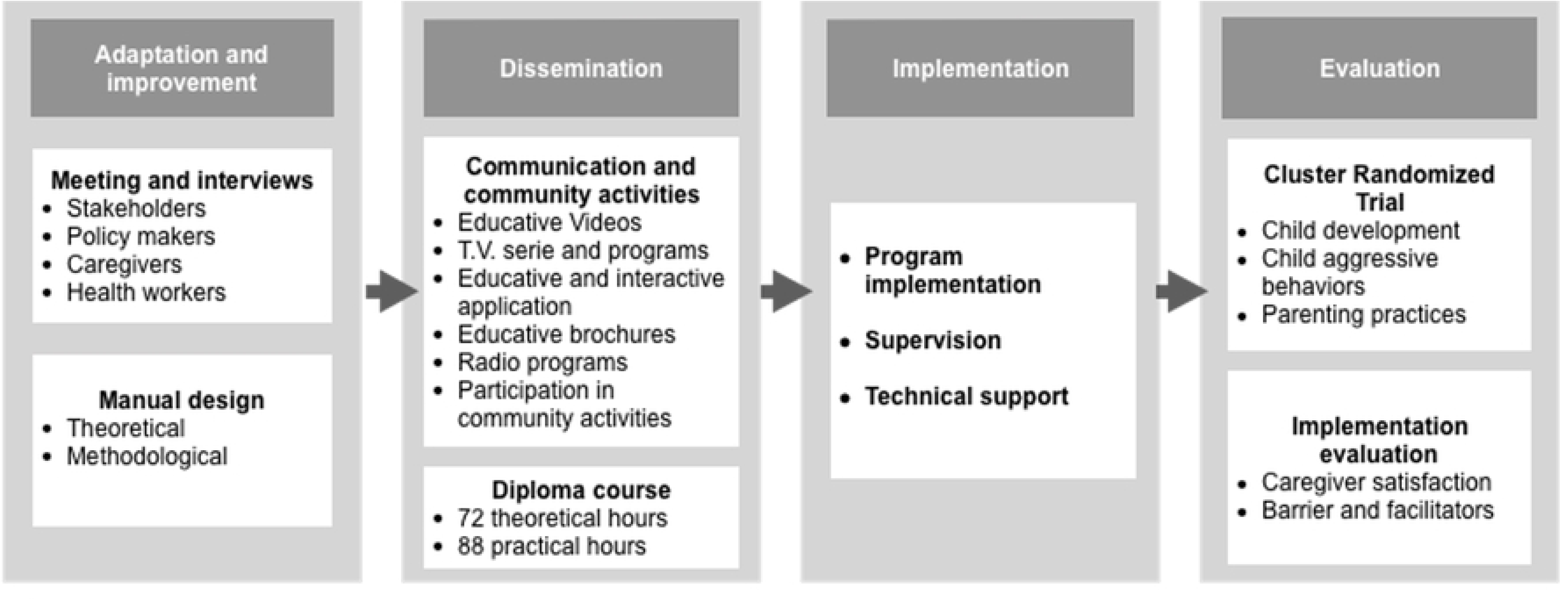

**Figure.**
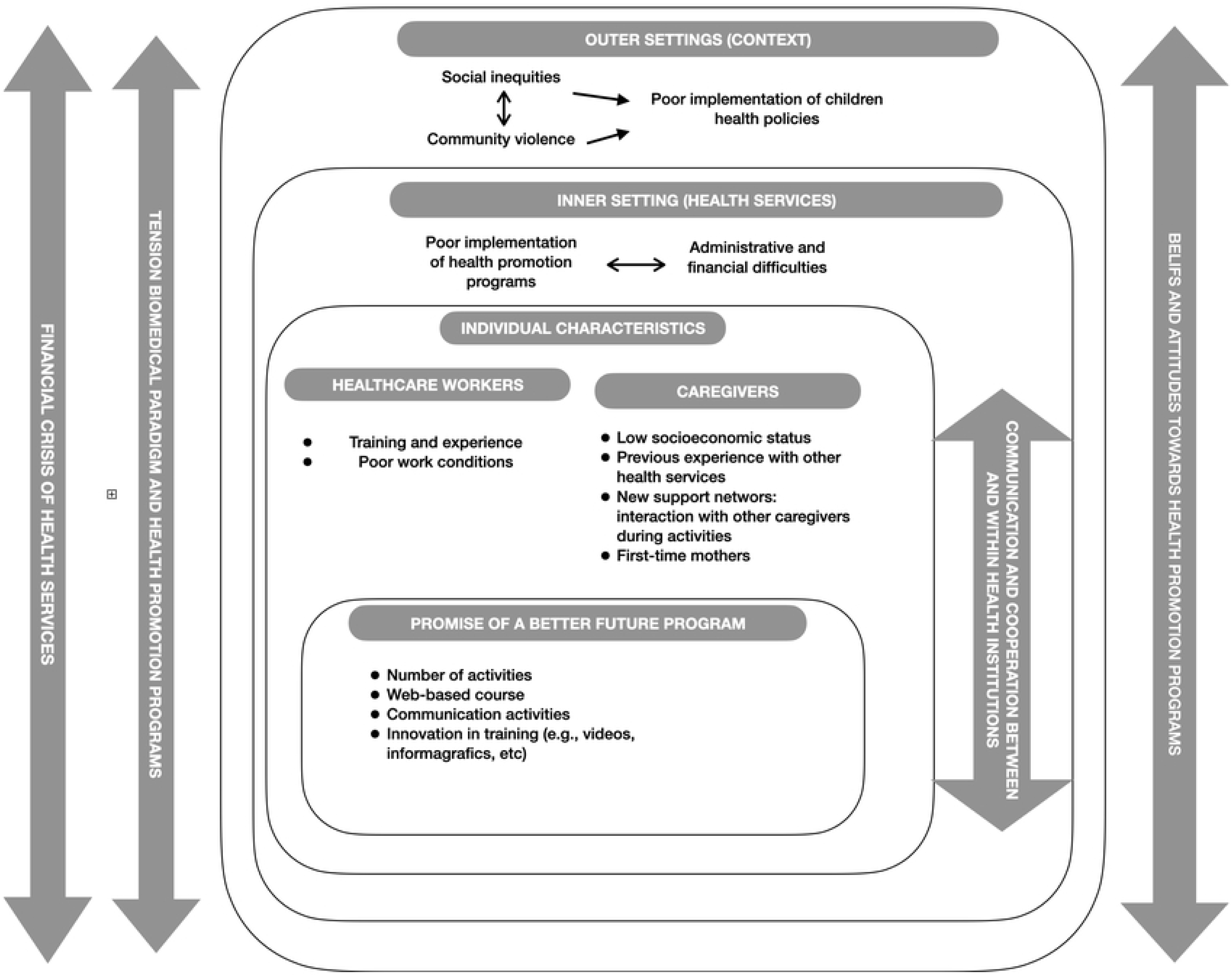

### Outer settings

#### Social inequalities

The program was implemented among low-income populations in a rural setting, and families have difficulty obtaining essential services for their children’s development (e.g., food, housing, clothing, education, and transportation). In general, the Grow and Development Programs connect families with other services and opportunities, and PVFP also supports families’ connections to these services. However, the program was not designed to address the basic needs directly. Therefore, some families were unable to attend or participate in the activities because their basic needs were not met, and parents had to work instead.

Community violence breeds mistrust. There are neighborhoods and municipalities with illegal armed groups related to drug trafficking, guerrillas, and paramilitary groups. This violence limited the caregivers’ and the professionals’ mobility to carry out the activities. Violence fosters general mistrust of others, particularly strangers. Some caregivers mistrust professionals and are reluctant to allow them to enter their homes, creating barriers to participation in program activities and to trusting healthcare workers’ proficiency.

> “Situations of violence that are occurring in the municipality, including the abduction of children, have generated alertness among mothers by decreasing the levels of assistance to the different programs that are being developed in the Hospital” (10 SWOT report).

#### Grade of Implementation of childhood and mental health public policies

Some municipalities have public policies that promote overall child development, including economic support, education, health, and family development. However, these policies are not fully implemented and are accompanied by additional barriers and facilitators. Some of these policies support growth and development programs, and a lack of continuity can directly affect the resources allocated to these programs, thereby impacting PBFP implementation. Also, the full implementation of public policies may address some of the social determinants of health. In municipalities that implemented public policies, the PBFP was readily implemented because additional personnel and funding were available.

#### Tension between the Biomedical paradigm Vs

*Health promotion and prevention approach*: There is a predominance of the biomedical paradigm in health services and the population. Health services are designed to diagnose and treat disease, but not to develop health promotion activities. Healthcare workers are trained to perform medical procedures but often lack sufficient training in health promotion activities. Due to this lack of training, health workers preferred not to conduct health promotion activities and limited their actions to providing caregivers with lists of appropriate and inappropriate behaviors. In addition, community members and caregivers placed greater importance on children’s diseases and treatments. Therefore, they primarily brought their children to health services only when they were ill.

*The financial crisis in Colombia’s health system:* the system is experiencing an economic crisis that undermines program implementation. The financial crisis led to the partial or complete closure of some health services, nonpayment to personnel, and a reduction in personnel. This financial crisis affects the quality of health services provided with limited resources.

Additionally, this crisis led to a decrease in personnel and an increase in demand for healthcare workers. Some Healthcare workers had to assume multiple functions to cover the population’s needs. As previously noted, these poor working conditions lead to dissatisfaction among healthcare workers.

#### Inner settings

*Implementation of health promotion and disease prevention programs:* PBFP implementation encountered fewer difficulties when conducted in health services with structured health promotion and disease prevention programs, which had more space, financial resources, personnel, and training. In addition, the administrative staff was more receptive to the implementation and understood the importance of this type of health program.

*Administrative and financial difficulties in health services:* Some health services experienced financial problems that affected hospital operations and ultimately influenced program implementation. For instance, closed services or mergers with other hospitals did not receive the insurance payments for the user’s healthcare services.

*Healthcare services leadership*: Some hospitals have difficulty planning program activities due to a scarcity of trained personnel. They also experience administrative staff instability. Hospital directors change frequently in response to political conditions in each municipality, and some program directors were replaced during program implementation. The constant changes in the leadership result in difficulties keeping the institution’s engagement in the program. Finally, some hospitals lacked adequate spaces for health promotion activities, such as clean, well-illuminated rooms. Therefore, the program’s activities were performed in small spaces that impeded interaction between caregivers. The inappropriateness of spaces was also highlighted in the caregiver satisfaction survey.

> “The delay in the election of the new hospital director has prevented the director from considering the program needs…” (Healthcare worker 2)

*Communication and cooperation between and within institutions* are essential for program success. Mayors’ offices and hospitals should share common goals with the activities they undertake in the community. When health workers have common goals and similar procedures, the program can maintain fidelity and quality. Cooperation can also reduce the proliferation of programs and activities aimed at promoting child development. In addition, health services sometimes provide repetitive or contradictory messages that confuse parents about caring for their children.

> “The lack of coordination between the hospital and the professionals in the social protection secretary who expresses their disagreement with the administrative procedures and work methodologies of the hospital” (8 SWOT report)

### Individuals’ characteristics

#### Healthcare workers

*Health workers’ training and experience:* Some health professionals lack sufficient training to implement health promotion programs. PBFP provided training that improved specific skills required for program implementation. Healthcare workers are usually trained to provide information about healthy behaviors that caregivers should follow. Still, they lack training in communication skills to share experiences with caregivers about parenting practices in a more horizontal relationship.

Satisfaction with working conditions: Some health professionals perceived health services as focused primarily on revenue generation and secondarily on patient care. Working conditions for healthcare workers have deteriorated amid the financial crisis in health services. They lack adequate income or stable employment. They were employed on short-term appointments; some were hired through intermediary cooperatives that offered lower salaries, and some health services experienced mass layoffs.

#### Caregivers

*Economic difficulties and transport:* The low socioeconomic status of the families disrupted adherence to the program. These families were unable to pay for transportation to participate in program activities and were absent from work because they had to attend activities or pay for childcare. As we observed these difficulties, we provided transportation assistance to some parents or conducted home visits.

*Build and learn from other caregivers:* Caregivers who have participated in group activities report learning from other caregivers’ experiences. Some parents built relationships outside the program, sharing experiences that helped them overcome parenting difficulties. Although caregivers learned from group experience, they appreciated the program’s flexibility in adapting to their needs, including home visits and the development of individualized activities.

*Caregiver’s previous experiences with the health services:* Caregivers who had negative experiences with the health services judged the program based on those experiences. They assumed that they would have the same problems and low quality as other health services. Therefore, caregivers preferred not to engage in the program or skipped activities. Program professionals and health workers had to make an additional effort to persuade them to attend program activities and initiatives.

*First-time mothers:* Mothers who had their first child showed greater motivation and interest in participating in the program because they felt it could address their multiple needs and questions. They could also learn from more experienced caregivers during group activities.

#### Program characteristics

*The number of activities*: We observed contradictory perceptions among parents regarding the number of activities. Some caregivers indicated that the program should offer fewer activities because they could not attend monthly sessions. However, some mothers requested additional activities.

*Web-based course and a combination of resources:* The abundance of videos and educational materials was essential for training professionals and engaging families. The caregiver’s videos provided an effortless means of transmitting information because participants could identify with the parent. Additionally, the program appeared on local television and radio.

These multiple communication activities promoted engagement and maternal participation, including videos, infographics, live history videos with parents, and WhatsApp reminders. *The program has strong adaptation and theoretical support*. Caregivers, professionals in the mayors’ offices, and health workers perceive the program as having strong theoretical support. They perceived strong credibility and acceptability. The solid theoretical support, extensive professional experience, and high-quality didactic materials enable caregivers and healthcare workers to adopt new practices. These positive beliefs and attitudes about the program increased program engagement and program adherence.

## Discussion

In the current analysis, we found that inequalities, violence in the communities, and a lack of implementation of the public policies about child development and mental health, as well as the presence of violence in the municipalities, were some of the contextual factors that impeded policy implementation. Violence was a significant obstacle to program delivery, undermining community trust in healthcare workers. We created strategies, such as home visiting, for those families unable to mobilize to attend the activities because of violence. However, violence is an ongoing risk for the health workers as they move through communities. PBFP also became a space in which caregivers and healthcare workers could talk about their previous traumatic experiences and address the consequences of community violence. Previous study conducted by Murphy et al, emphasized the importance of increasing the evidence about ECDP implementation in emergency settings because this evidence could increase the quality of services created to protect children from the multiple negative consequences of conflict (2, 14).

Regarding service characteristics, we identified difficulties in healthcare services leadership that hindered administration and planning, thereby affecting resource allocation within the program. Inadequate implementation of health promotion programs directly impacts the delivery of PBFP. Difficulties with planning and funding could reduce the effective use of resources. In addition, a lack of leadership leads healthcare workers to adopt various procedures and goal-setting strategies that undermine intervention fidelity. Grol also described leadership as an important factor to implement evidence-based practices in health services, as applied to diabetes prevention (19).

Regarding provider characteristics, we found that poor working conditions and a lack of training in health-promotion activities were barriers to PBFP implementation. A systematic review reported a 40% prevalence of job exhaustion and a 19% prevalence of low personal accomplishment among health professionals (20). Previous research among nurses has shown that high job stress, burnout syndrome, and job dissatisfaction can affect service quality (21). In a study of general internists, low job satisfaction was associated with patients’ dissatisfaction with their health care. Additionally, previous research indicated that providing training to health workers may increase their job satisfaction by enhancing their professional development (22). Developing a certificate training program for healthcare workers could have positively affected their engagement and alleviated some of the job dissatisfaction.

We found that caregivers were highly satisfied with the PBFP activities; however, prior experience with the health service limited their engagement in the program. The research of MCHNP reported that previous experience with the health services determines the mother’s participation in the program (11). Finally, we described how first-time mothers were more interested in and engaged with the program than mothers with more than one child. According to the transtheoretical model of change, the first step toward change entails motivation and recognition of the necessity of change (23). As a result, some first-time mothers improved more during the intervention than did mothers with multiple children. Since first-time mothers are more engaged and satisfied with the program, PBFP may be effective at engaging and changing the behaviors in first-time mothers (17).

Regarding program characteristics, while some caregivers perceived PBFP as having numerous activities, others felt they could benefit from attending more sessions. The difference can be explained by the fact that some mothers had more time to attend or greater parenting-related stress. Mothers who expressed interest in the program reported numerous barriers to participation, including limited time to attend and a lack of funds for childcare. Communication activities were valued for their educational value and for fostering engagement and connection with the program.

In this analysis, we identified several cross-cutting constructs that affect program implementation at multiple levels, including the predominance of the biomedical model over a health promotion and prevention approach, the financial crisis in health services, beliefs and attitudes toward the program, and communication and cooperation among institutions.

The biomedical model emphasizes the detection and treatment of disease but places insufficient emphasis on health promotion and disease prevention. As a result, programs like PBFP are not prioritized by stakeholders, health services, professionals, or families. This emphasis on the biomedical model fragments services into those for disease and those for prevention, even though the health-disease process is continuous in the individuals. The predominance of the biomedical model over health promotion has been described in previous qualitative evaluations of growth and development programs in Colombia (24). Although the program provided training in communication skills, it is difficult to change the paradigm of healthcare systems and culture. Therefore, we believe it is necessary to revise curricula in the healthcare profession by emphasizing not only biomedical knowledge but also the ability to deliver health promotion services and health education. Hays et al. studied the inclusion of health promotion in physician training. He found that curricula increased health-promotion content. However, most of the evidence that he presented belongs to High Income Countries (HIC), it is necessary to continue updating health promotion topics in the curricula in LMIC (25).

The financial crisis in the health system affects the quality of health services and the working conditions of health workers. The economic crisis was an essential determinant of PBFP implementation, affecting different aspects of quality and resources, health services, and job satisfaction of health workers, and perceived quality of health services by caregivers. Other publications and newspapers have described the financial crisis in Colombia’s health system (26, 27). We observed that the financial crisis not only affects current services but may also affect innovations implemented within those services. It has profound implications for the future and quality of health services, as these services may not keep pace with technological and scientific advances. This aspect can have important implications for the implementation of programs in LMICs, which may also face financial challenges in health services.

Planning and financial programs reduced the time required and the degree of health professionals’ dedication to the program. Even though we signed agreements with the hospital and major offices, it remained challenging to sustain professionals’ commitment to the training and activities. We had to meet multiple times with staff and healthcare workers to reschedule the activities. The delivery of PBFP was easier in health services with strong primary care, as healthcare workers were more receptive than those in services with limited health promotion programs. According to the transtheoretical model of change, professionals with prior training in health promotion may be more predisposed to change their practices than those without such training.

We emphasize the program’s impact on caregivers’ support networks. This positive and unexpected effect enables caregivers to establish relationships and foster collaboration outside the program setting, thereby significantly benefiting families by creating new support networks. Social networks are essential because they enhance caregivers’ mental health, enable them to provide better care for the child, and ultimately support the child’s development (28).

Among the limitations of this study, we note that we did not include interviews with policymakers in the PBFP program. However, they participated in the monthly meetings to discuss the program’s strengths and barriers. In addition, we need to assess in the future how communication activities, such as videos and infographics, have improved PBFP implementation.

Results from this research can be used not only to improve the implementation of the PBFP but also to provide evidence to guide the implementation of similar health services in LMICs and countries with a history of violence. Results may also provide evidence on how conflict affects access to programs that mitigate the mental health and health service consequences of conflict (29).

## Conclusion

Poor family conditions, financial stability of the health system, cooperation among health institutions, and the beliefs and attitudes of different actors toward the program are important aspects to address when implementing an early child development program. Additionally, appropriate conditions for healthcare workers can determine the success of a program. Finally, caregiver characteristics, such as being a first-time mother and trust in health services, can be important facilitators of program implementation.

## Data Availability

The datasets used and/or analyzed during the current study are available from the corresponding author by request.

## Acknowledgements

We thank the caregivers, children, municipalities, health institutions, and healthcare workers. Also, I thank Sarah Ayton for review of the English grammar and style.

## Author’s contributions

ARH conceived the study, supervised the study, validated the study, visualized the study, contributed to the conceptualization and data curation, participated in qualitative and quantitative analyses, and was responsible for drafting and finalizing the manuscript. MPAM supports writing and analysis. JWCO was responsible for funding acquisition. JWCO and JOC participated in writing, methodology, and analysis. AMC collected interview and participant information for the qualitative study. NEMG was responsible for conceptualization, writing, data collection, and quantitative data analysis. All authors read the manuscript and approved the final manuscript version.

## Ethics Statement

This project was approved by the bioethics committee at the National School of Public Health at the University of Antioquia, Colombia (Act 125, August 6 of 2025). All participants (parents and healthcare workers) read and signed informed consent. Caregivers and healthcare workers were informed about the risks and benefits of participating in the research, as well as their anonymity and rights.

